# Assessment of Motion Bias on the Detection of Dopamine Response to Challenge

**DOI:** 10.1101/2021.02.18.21252006

**Authors:** Michael A. Levine, Finnegan Calabro, David Izquierdo-Garcia, Daniel B. Chonde, Kevin T. Chen, Inki Hong, Julie C. Price, Beatriz Luna, Ciprian Catana

**Affiliations:** Athinoula A. Martinos Center for Biomedical Imaging, Department of Radiology, Massachusetts General Hospital and Harvard Medical School, Charlestown, Massachusetts; Department of Psychiatry, University of Pittsburgh Medical Center, Pittsburgh, Pennsylvania; Department of Bioengineering, University of Pittsburgh, Pittsburgh Pennsylvania; Siemens Healthcare MI, Knoxville, Tennessee

**Author notes:** Corresponding author: Ciprian Catana, A.A. Martinos Center, MGH, 149 13^th^ St. Charlestown MA, 02129. Disclaimer: none. First author: Michael A. Levine, A.A. Martinos Center, MGH, 149 13^th^ St. Charlestown MA, 02129.

**Keywords:** PET, ^11^C-Raclopride, behavioral task, kinetic modeling, motion correction.

## Abstract

^11^C-Raclopride (RAC) positron emission tomography (PET) is used to study dopamine response to pharmacological and behavioral challenges. Behavioral challenges produce smaller responses than pharmacological challenges and are more susceptible to sources of bias, including motion bias. The purpose of this study was to characterize the effect of motion bias within the context of a behavioral task challenge, examining the impact of different motion correction strategies, different task response magnitudes, and intra-versus interframe motion.

**Methods:** Seventy healthy young adults were administered bolus plus constant infusion ^11^C-Raclopride (RAC) and imaged for 90 min on a 3-Tesla simultaneous PET/magnetic resonance (MR) scanner during which a functional MRI (fMRI) reward task experiment was conducted. Kinetic analysis was performed using an extension of the multilinear reference tissue model (MRTM), which encoded the task response as a unit step function at the start of the task (t = 40 min). The quantitative impacts of different approaches to motion correction (frame-based, reconstruction-based, none) were compared using voxel maps of change in binding potential (ΔBP_ND_). Motion bias was compared to task effect by simulating different levels of ΔBP_ND_ (0%, 5%, 10%, 20%) in conjunction with simulating high and no motion. Intraframe motion was simulated using motion estimates derived from the simultaneously acquired MR data. The relative impact of intraframe motion was evaluated by comparing maps of bias in ΔBP_ND_ before and after applying frame-based motion correction.

**Results:** Among the high-motion subjects, failure to perform motion correction resulted in large artifacts. Frame- and reconstruction-based approaches both corrected for motion effectively, with the former showing moderately more intense ΔBP_ND_ values (both positive and negative) in and around the striatum. At low task response magnitudes, simulations showed that motion bias can have a greater relative effect. At 5% ΔBP_ND_, motion bias accounted for 60% of the total bias, while at 10% ΔBP_ND_, it accounted for only 34%. Simulating high-temporal resolution motion, frame-based motion correction was shown to counteract the majority of the of the motion bias effect. The remaining bias attributable to intraframe motion accounted for only 8% of the total.

**Conclusion:** Motion bias can have a corrupting effect on RAC studies of behavioral task challenges, particularly as the magnitude of the response decreases. Applying motion correction mitigates most of the bias, and specifically correcting for interframe motion provides the bulk of the benefit.

## INTRODUCTION

^11^C-Raclopride (RAC) positron emission tomography (PET) is a well-established means of imaging dopamine D_2_/D_3_ receptors. It has been used to measure the spatial distribution of receptors^1^, as well as the kinetic properties of their binding^2^. It has also been used to study the dopaminergic system under blocking and displacement conditions and after pharmacological challenges such as amphetamine^3-5^, cocaine^6, 7^, and nicotine^8, 9^, and behavioral challenges such as reward^10, 11^, motor performance^12^, pain^13^, and cognitive tasks^14^.

One of the difficulties in working with behavioral challenge experiments is their modest response magnitudes relative to those of pharmacological challenges. While pharmacological challenges may cause changes in binding potential (BP_ND_) of 10-20% or greater^5, 15, 16^, behavioral challenges produce smaller changes of around 0-10%^14, 17-19^. With these smaller effects, measurements of behavioral challenge response are more difficult to discern from noise, while also being more susceptible to different sources of bias^20, 21^, including bias arising from head motion^17^.

Motion bias is routinely addressed in the field using either frame-based motion correction^22, 23^ or more advanced methods which require independently estimated motion (e.g. reconstruction-based motion correction^24^ or event-based rebinning^25, 26^). In this work, we build upon an earlier finding of measurable differences in ΔBP_ND_ between participant strata^27^ to investigate motion bias for participants with different motion magnitudes, using different motion correction techniques, and relative to different challenge response sizes. While others have demonstrated the effect of motion on PET quantification^28-30^ and with RAC in particular^31-33^, we characterized the effect of motion bias specifically on measurements of intra-scan behavioral challenge.

First, we evaluated the impact of motion bias on behavioral challenge response as measured by voxel maps of change in binding potential (ΔBP_ND_). To maximize our power to observe its effect, we contrasted the impact of motion bias in the subgroups of participants with the highest motion and the lowest motion. Within these participant groups, we compared three different approaches to motion correction: no motion correction (maximizing motion bias), frame-based motion correction (the standard in the field), and a reconstruction-based method that uses a data-driven approach to estimating motion (greater complexity and temporal resolution). Then, in order to conceptually disentangle the behavioral task response from motion bias, we used simulations to impose different magnitudes of task response and compared the impact of motion bias to the simulated ground truth at each level of response. Finally, we used high temporal resolution MR-based motion estimates to simulate intraframe motion. By applying standard frame-based motion correction to this simulated data with simulated motion, we were able to distinguish between the biasing effects of inter- and intraframe motion together (before motion correction) and intraframe motion alone (after motion correction).

## MATERIALS AND METHODS

### Participants and Data Acquisition Protocols

70 adult participants (female = 34, ages 18-30 years) were retrospectively selected from an adolescent development study performed on an integrated PET/MRI scanner (Biograph mMR, Siemens Healthineers, Erlangen, Germany) at the University of Pittsburgh Medical Center^34^. Healthy volunteers were scanned using RAC while simultaneously performing a functional magnetic resonance imaging (fMRI) task designed to study reward response. All participants gave written informed consent and were studied in accordance with experimental procedures approved by the University of Pittsburgh Institutional Review Board. All participants were imaged in the headfirst supine position and PET data were collected from the start of radiotracer administration.

Participants received a bolus injection of RAC (661-802 MBq) followed by a constant infusion (KBol = 105 min) for the duration of the 90-minute scan. PET 3D coincidence event data were collected and stored in list-mode format. PET volumes were iteratively reconstructed with a uniform 3-minute framing using the e7 tools (OP-OSEM, 3 iterations, 21 subsets, no filter)^35, 36^. Attenuation correction was performed using a pseudoCT attenuation map^37^ generated from a T1-weighted structural sequence (MPRAGE) registered to the PET volume at the start of the task. The simultaneously acquired MRI protocol centered on eight fMRI-BOLD sequences (echo time [TE] = 30 ms, repetition time [TR] = 1500 ms). Six were acquired during the task window and two were resting state sequences, one at the beginning of the scan and one after completion of the task. The protocol also included an MPRAGE for attenuation and region-of-interest label generation, and several sequences used routinely for brain studies (e.g. localizer, gradient-echo field mapping, ultrashort TE, TurboFLASH, diffusion spectrum imaging, and magnetization transfer ratio). Six participants were censored from the analysis, one because the imaging study was interrupted, and five because the PET scan ended before the last resting state fMRI sequence was completed. Further information about the study design can be found in the primary reports^27, 34^.

### Motion Correction

Two different approaches for addressing participant motion were compared: frame-based motion correction and reconstruction-based motion correction.

#### Frame-based Motion Correction

The methodology for frame-based motion correction is well established in the field^22, 23, 38, 39^. Briefly, the head was treated as non-deformable, and a linear least squares rigid body registration algorithm (implemented in SPM8^40^) was applied to track its displacements. This was accomplished by retrospectively coregistering the dynamic volumes^41^ to a reference volume at the onset of the task (t = 40 min).

#### Reconstruction-based Motion Correction

To correct for motion during reconstruction, it was first estimated from the PET data using a procedure similar to frame-based motion correction. First, the PET list-mode data were split into 1-minute subframes (with the exception of the first subframe, which was 2 minutes to accommodate the rapidly changing spatial contrast associated with delivery). Next, preliminary image reconstructions were performed, accounting for detector normalization and radiofrequency MRI coil array attenuation, but without correcting for head attenuation. The resulting volumes were registered to produce motion estimates, rather than to perform motion correction directly. These motion estimates were measured as a set of three translations and three rotations (e.g. **Figure 2A**), which were then encoded into a 4×4 transformation matrix and supplied to the reconstruction algorithm for motion correction.

Motion was corrected as part of the iterative PET reconstruction for each dynamic frame using a development version of the e7 tools^24^. For each PET volume reconstructed, the associated timeframe was divided into subframes corresponding to the temporal resolution of the motion estimates. During each reconstruction iteration, the current estimate of the PET volume was replicated for each subframe, and the subframe volumes were moved according to the estimated motion before being forward projected into sinogram space. After backprojection, the subframe volumes were motion corrected and averaged to form the subsequent estimate of the reconstructed PET volume. Correction for scatter and randoms was performed using the full sinogram for the given timeframe, with proportional scaling for each subframe.

### Measuring Total Motion

To quantify the total extent of motion estimated from the PET data, an ‘aggregate distance’ metric was calculated by comparing how a 3D mesh of points evenly distributed throughout the participant’s head would be displaced by the estimated motion. For each of the points in the mesh, the distance from its original position to its motion-displaced position (as estimated by the PET registration) was calculated. These distances were then averaged across all points to create the aggregate distance metric at every timepoint throughout the scan. Cumulative motion was calculated by summing the aggregate distance for all timepoints.

### Quantitative Impact of Motion Correction

To demonstrate the negative impact of motion and the compensatory effects of frame-based and reconstruction-based motion correction, a comparison was performed between participants who moved the most and those who moved the least. The high and low motion groups were selected to be the top and bottom 20% of participants (N = 10 per) as measured by the total cumulative motion (**Figure 1**).

**Figure 1:**
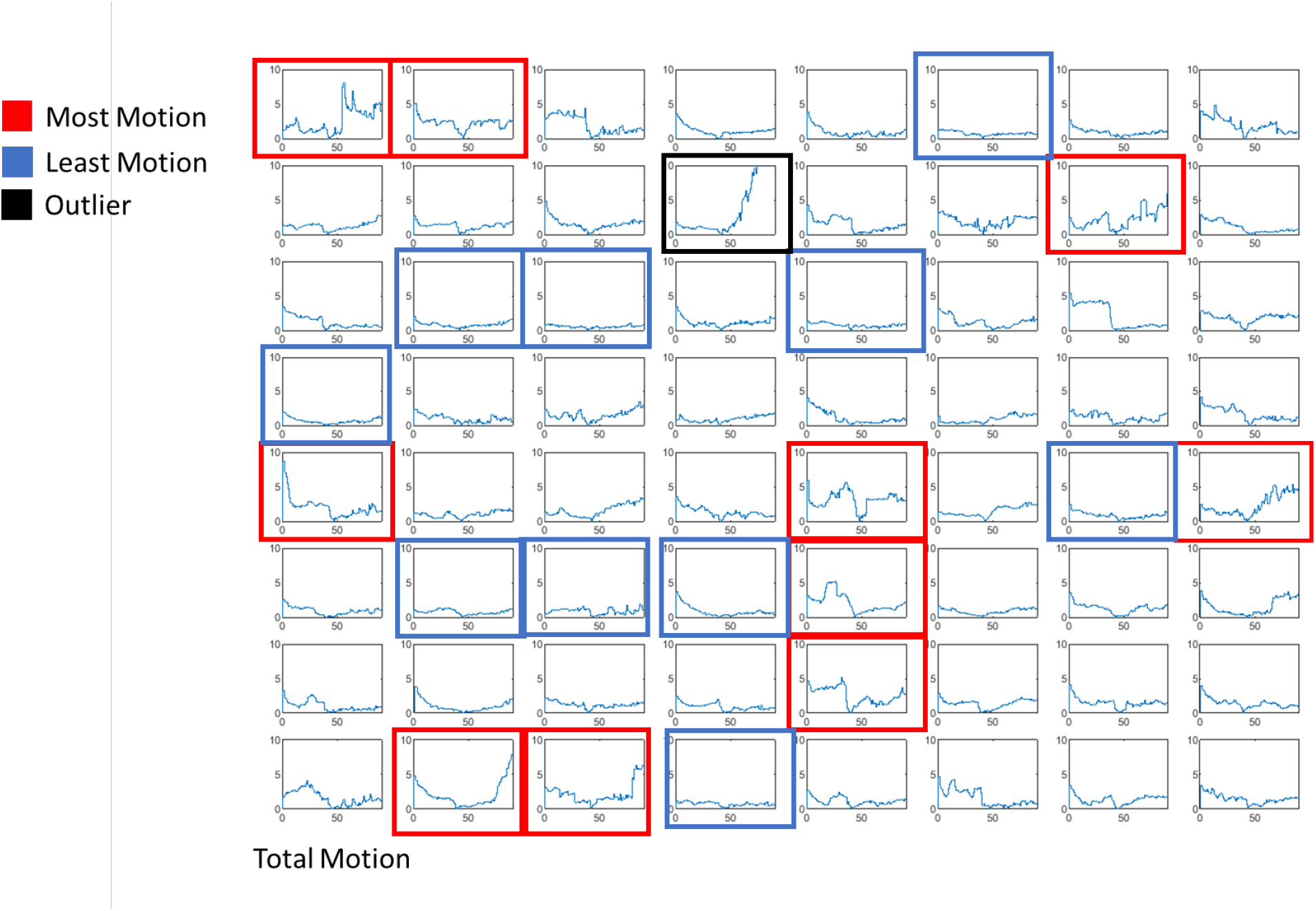
Participant Total Motion. Each subplot represents a single participant, with total motion (mm) plotted against time post injection (min). High (red boxes) and low (blue boxes) motion groups were created by selecting the top and bottom 20% of participants by total motion.

**Figure 2:**
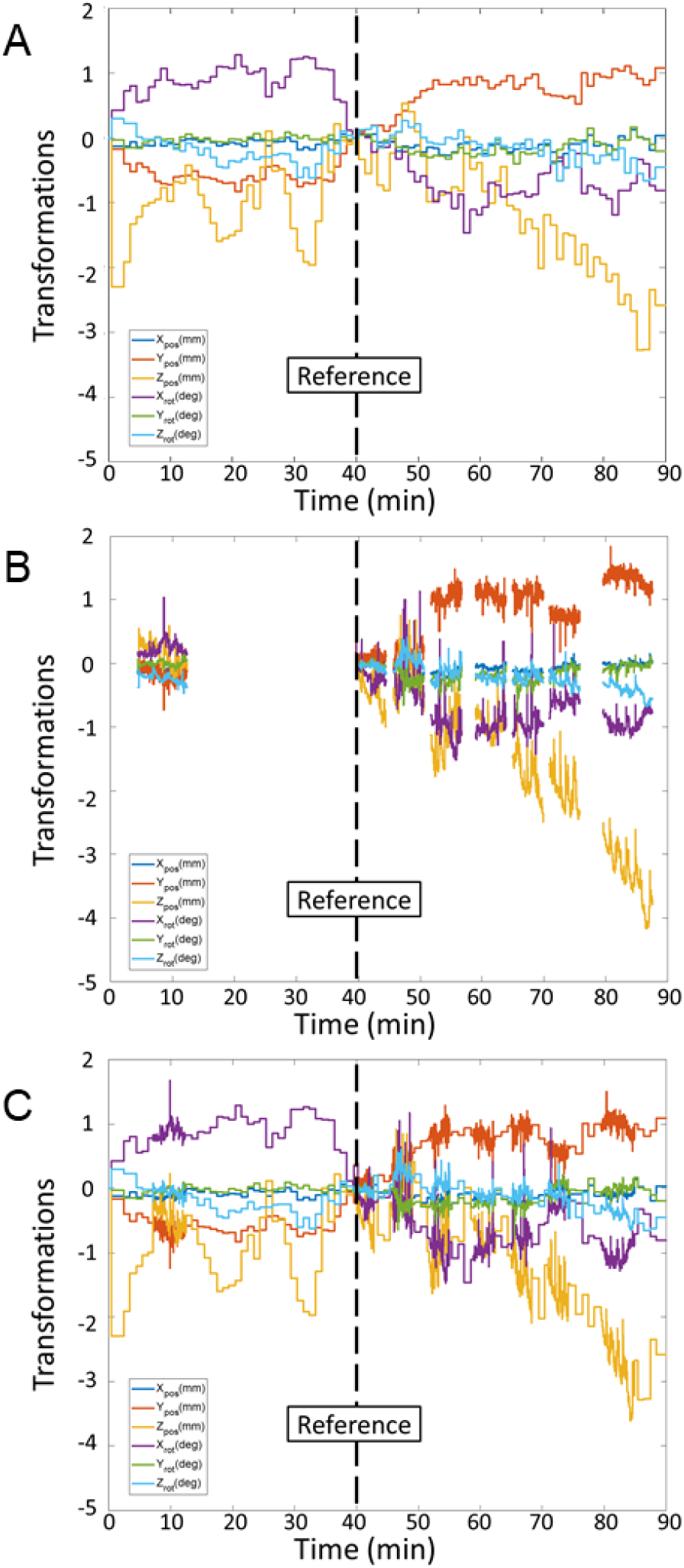
Example PET, MR and Unified Motion Estimates. (A) PET based estimates with temporal resolution 1 min and reference t = 40 min. (B) MR-based estimates with gaps between sequences, temporal resolution 1.5 sec, reference t = 40 min. (C) Unified motion estimates with PET estimates replaced with higher resolution MR estimates where available.

For each participant, voxel maps of binding potential and the change in binding potential (ΔBP_ND_, i.e., BP_ND-POST_ – BP_ND_) were generated using kinetic modeling of the data obtained from the images motion corrected using the methods described. The Multilinear Reference Tissue Model (MRTM)^42^ was used to measure the BP_ND_ in the striatum (where dopamine receptors are most abundant) with the cerebellum (negligible dopamine D_2_/D_3_ receptor concentration) as the reference tissue (Eq. 1). When considering a model with a single tissue compartment, BP_ND_ is expressed in terms of a combination of the true efflux rate (k_2_) and the apparent efflux rate (k_2a_)^43^ (Eq. 2).

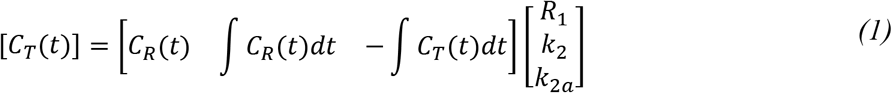

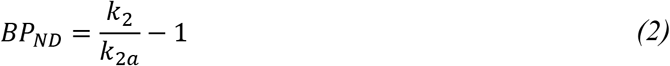

The model was then extended (Eq. 3) to accommodate a behavioral challenge using the formulation of Alpert et al.^44^ with a unit step at the onset of the task (t = 40 min) as the activation function. This is similar to the approach taken by Normandin et al.^45^, though with a simpler challenge encoding.

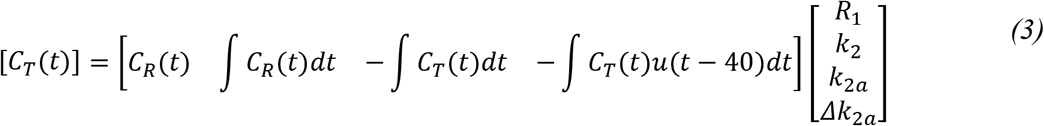

In order to decrease the effect of noise, the model was reduced to an extension of MRTM2 (Eq. *4*) by fixing 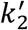 to the value obtained from a first pass fit of a high binding region (putamen)^42,46.^

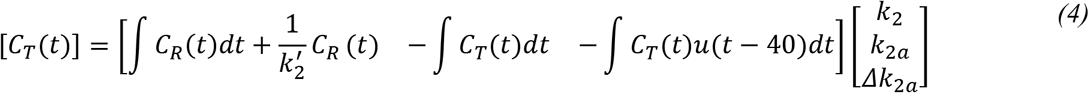

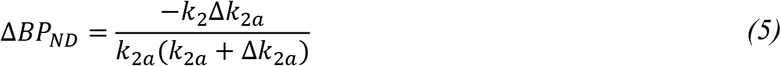

Fitting the model on a voxelwise basis produced parametric maps of BP_ND_ (Eq. 2) and ΔBP_ND_ (Eq. 5), which were spatially normalized to the Montreal Neurological Institute template using FreeSurfer’s combined volumetric and surface registration^47^ and averaged across participants within the high and low motion groups.

### Simulating Motion Bias

Simulations were performed to evaluate the effect of motion bias on task responses with different assigned magnitudes. To simulate full voxelwise PET timeseries data, individual microparameter (*k*_1_, *k*_2_, *k*_3_, *k*_4_) values for every voxel were required. Voxel maps of R_1_ and BP_ND_ were obtained by averaging MRTM fit values across study participants. Global values of 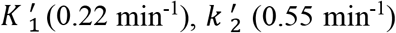, and *k*_4_ (0.13 min^-1^) were set according to nonlinear fits from a different RAC study which included arterial blood plasma draws^5^. These were used together according to the following equations (Eqs. 6-8)^43^ to obtain microparameters for all voxels.

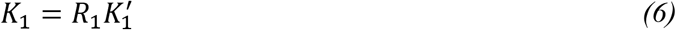

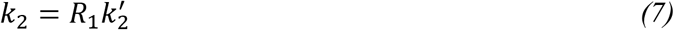

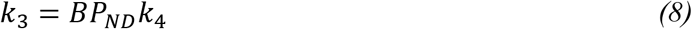

Although RAC data is best represented by a two-tissue compartmental model (2TCM)^2, 48^, a one tissue compartmental model (1TCM) was used for the simulation, conforming to the 1TCM topology of the model used for fitting (an extension of MRTM2), and thereby avoiding confounding from model bias. Therefore, each voxel time activity curve (TAC) was simulated using only two microparameters (*k*_1_, *k*_2*a*_), instead of four (*k*_1_, *k*_2_, *k*_3_, *k*_4_)^43^.

The magnitude of a simulated challenge response (Δ*BP_ND_*) within the striatum was varied (0%,-5%, −10%, −20%) to assess the impact of motion bias at different levels of challenge response. The arterial input function was simulated as a bolus plus constant infusion, with the bolus modeled as a gamma variate function^49^, and the constant infusion rate set according to that of the study (K_Bol_ = 105 min). The specific input function was selected from this family by determining the curve that, when paired with the chosen rate parameters, created reference (cerebellum) and target (putamen) TACs that most closely matched the acquired data. Motion was added to these simulations by transforming each reconstructed PET volume by a set of motion parameters. Different motion realizations were simulated by using motion parameters estimated from each participant in the high motion group.

These simulated PET volumes were analyzed in the same manner as the acquired data: kinetically modeled using an extension of MRTM2 that adds a unit step at the start of the task. The parametric maps from the high motion simulations were averaged across motion realizations. The analyses of the motionless and high motion simulations were compared using the parametric maps of Δ*BP_ND_*. Bias was assessed by evaluating the absolute value of error in ΔBP_ND_ relative to the simulated ground truth at the voxel level. Bias attributable specifically to motion was isolated by comparing the absolute error maps between the high motion and no motion simulations.

### Simulating Intraframe Motion

To simulate realistic motion at a high temporal resolution, motion was estimated from the simultaneously acquired MR data. The algorithm for estimating motion from EPI-based MRI sequences has been described previously^25, 50^. It follows the same approach as the PET-based approach (**Figure 2A**), using rigid body registration with a least squares cost function to estimate motion. The first volume in the first task fMRI sequence was selected as the reference volume, and all other volumes were registered to it in two steps: intra-sequence and inter-sequence. First, volumes within the same sequence were registered, producing a set of motion estimates for each volume. Afterward, the reference volume used for each sequence was registered to the overall reference volume at the start of the task. Final motion estimates were obtained by multiplying the 4×4 transformation matrices corresponding to the intra- and inter-sequence estimates (**Figure 2B**).

Higher temporal resolution estimates were created by substituting the PET-for MR-based estimates wherever they were available, creating unified motion estimates. However, to avoid discontinuities within the unified motion estimates, instead of using intersequence MR-based estimates for the second transformation, the time-matched PET-based transforms were used (**Figure 2C**).

PET volumes were simulated with a dynamic framing matching the high temporal resolution of these motion estimates and transformed according to them to create intraframe motion. To create a series of volumes with a uniform framing of 3 minutes, the subframe volumes at this higher temporal resolution were averaged. Where motion is simulated between the subframes, the averaging creates a blurring effect in the final volume, simulating intraframe motion.

The relative effects of intra- and interframe motion were compared by simulating intraframe motion with no task challenge and either performing no motion correction or frame-based motion correction before kinetic analysis. Not performing motion correction allowed us to estimate the total motion bias in ΔBP_ND_, while correcting for it allowed us to estimate the contribution of the intraframe component alone.

## RESULTS

### Quantitative Impact of Motion Correction

Total motion over the course of the scan was plotted for all study participants (**Figure 1**), showing an average cumulative distance travelled of 4.6 mm (range 0.9 mm – 16.6 mm). The total motion was used to group participants, selecting the top and bottom quintiles to create high and low motion groups which differed substantially in their degree of motion corruption and bias (**Figure 3**). One participant was excluded from the grouping due to extreme motion that could not be sufficiently corrected. While cases with low motion benefit minimally from motion correction, participants in the high motion group saw improvements in the reconstructed PET data and kinetic model fits. Example regional TACs demonstrate the impact of motion correction (**Figure 4**). The cerebellum is a large enough region (∼140K voxels) that its TACs are robust to motion corruption, while the TACs in the smaller nucleus accumbens (∼1.4K voxels) are much more susceptible, as seen in the high motion group (top row). To assess the quality of the kinetic model fits, we measured the Akaike Information Criterion (AIC)^51^ and used it to compare across motion correction approaches with a paired t-test. In the high motion group, significant decreases in the AIC were observed in the nucleus accumbens after motion correction (**Figure 5**).

**Figure 3:**
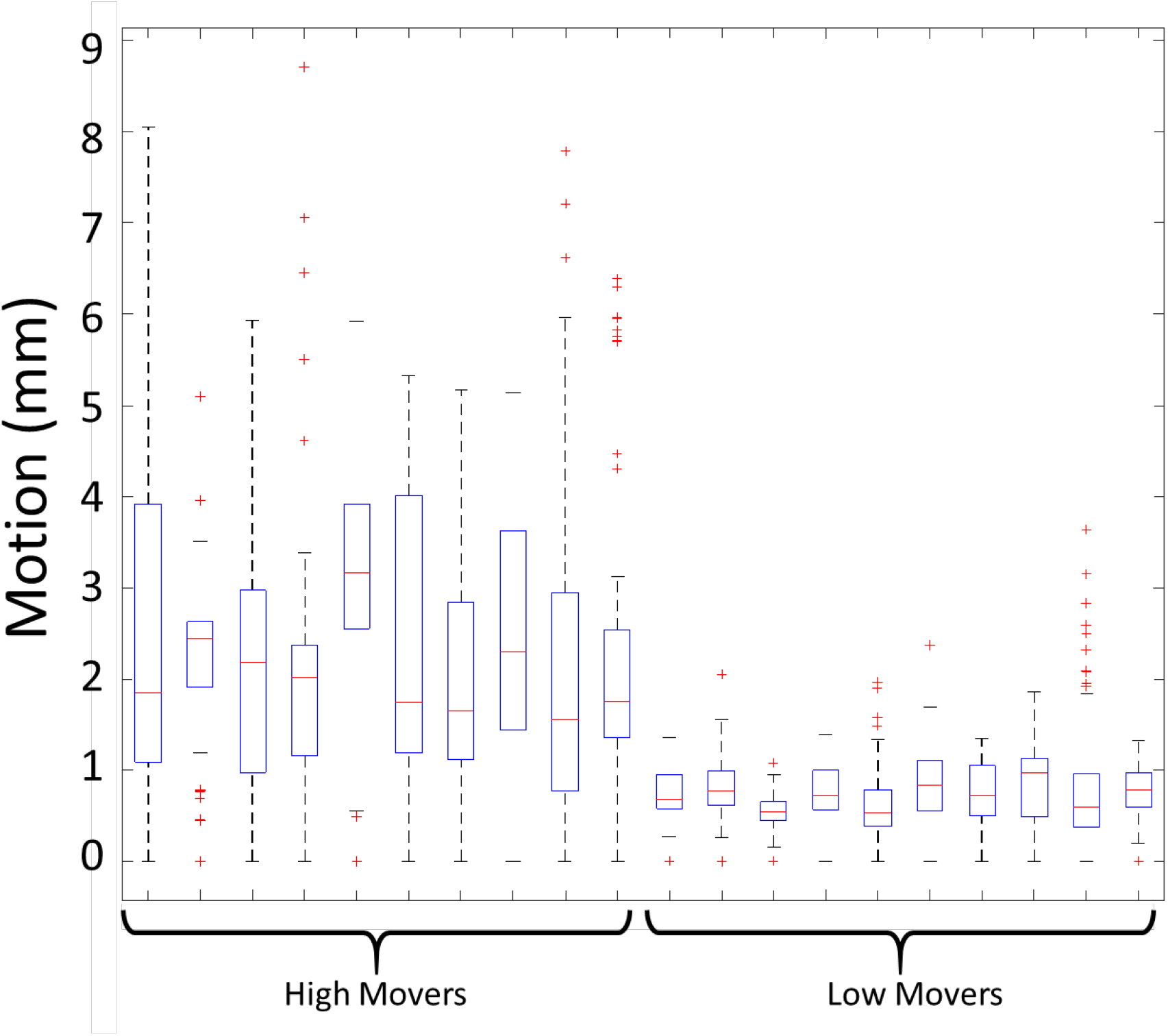
Differences in total motion between high motion and low motion groups. Each box represents the distribution of an individual participant’s motion timecourse (as shown in **Figure 1**).

**Figure 4:**
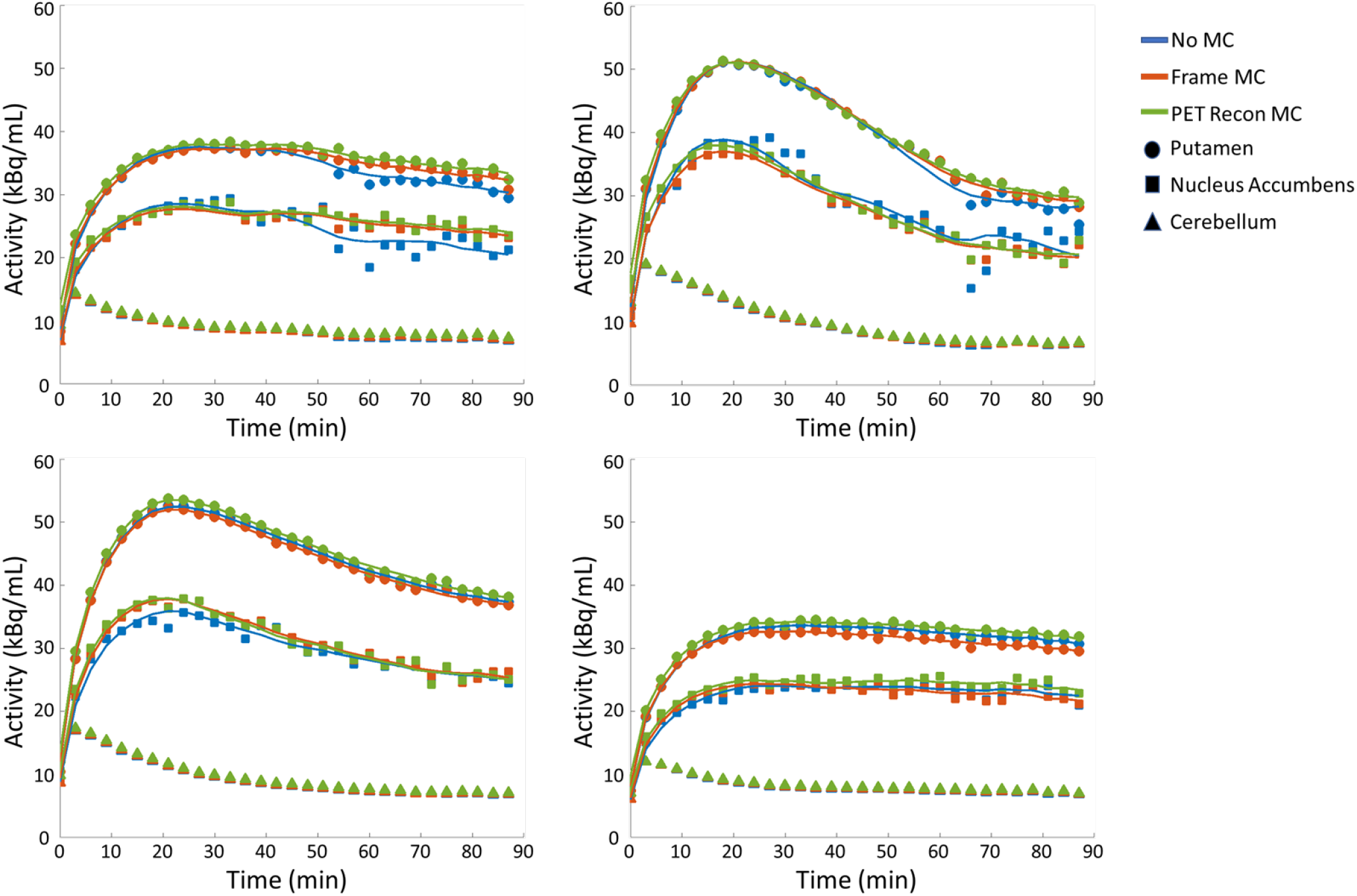
Example Time Activity Curves and kinetic model fits. Top Row: Representative participants from the high motion group. Bottom Row: Representative participants from the low motion group.

**Figure 5:**
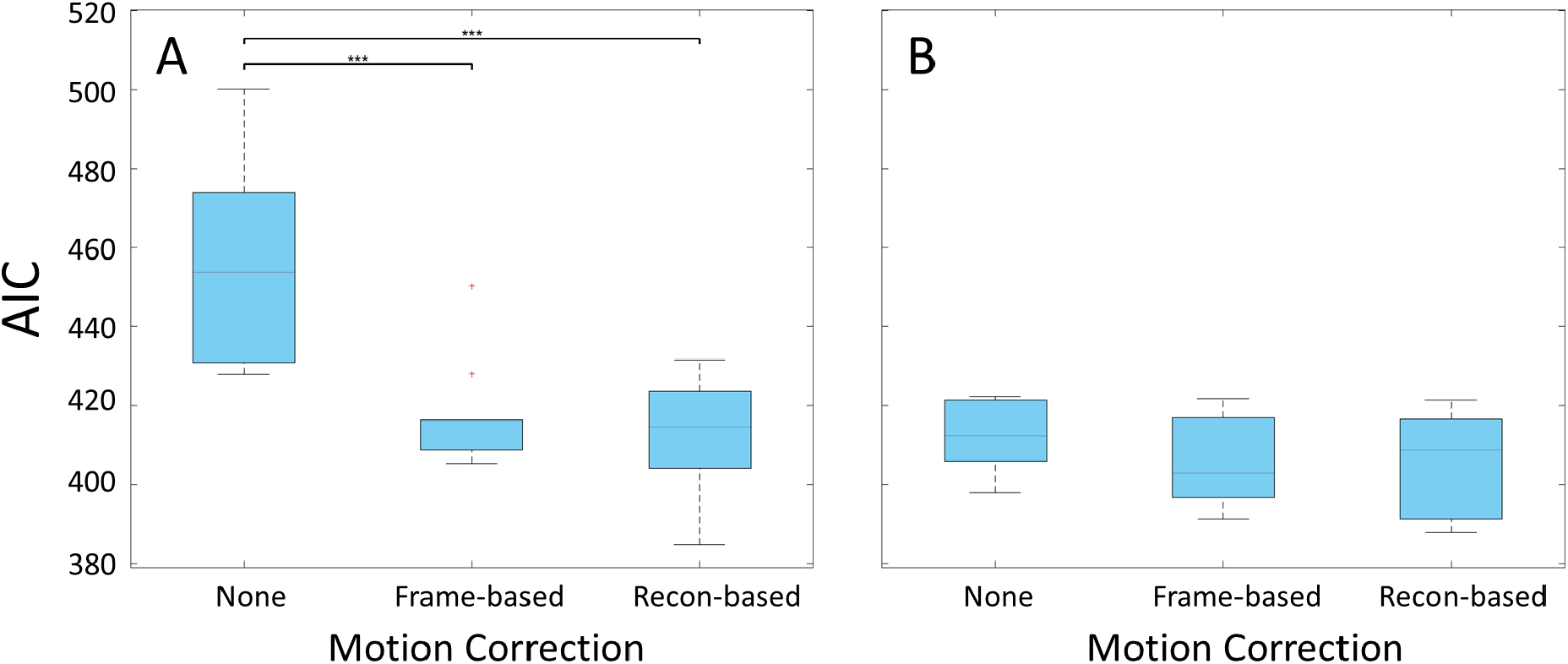
Akaike Information Criterion after kinetic modeling in the nucleus accumbens. (A) Participants in the high motion group. (B) Participants in the low motion group. (*** p < 0.001)

Figure 6. shows voxel maps of task response (ΔBP_ND_) in high and low motion groups using different motion correction schemes. In the case where motion was high, lack of correction resulted in large positive and negative biases around the striatum, while frame-based motion correction removed most of the bias and reconstruction-based motion correction eliminated even more. When averaging these values within the putamen—the highest binding region—no motion correction resulted in the greatest change in binding potential (−10.7%), while frame-based (−2.9%) and reconstruction-based (−0.5%) correction showed smaller ΔBP_ND_ values (**Table 1**). The low motion group, in contrast, was far less affected by the motion correction approach used, yielding ΔBP_ND_ values of −2.7%, −2.0%, and −0.3% for no motion correction, frame-based motion correction, and reconstruction-based motion correction respectively (Table 1).

**Figure 6:**
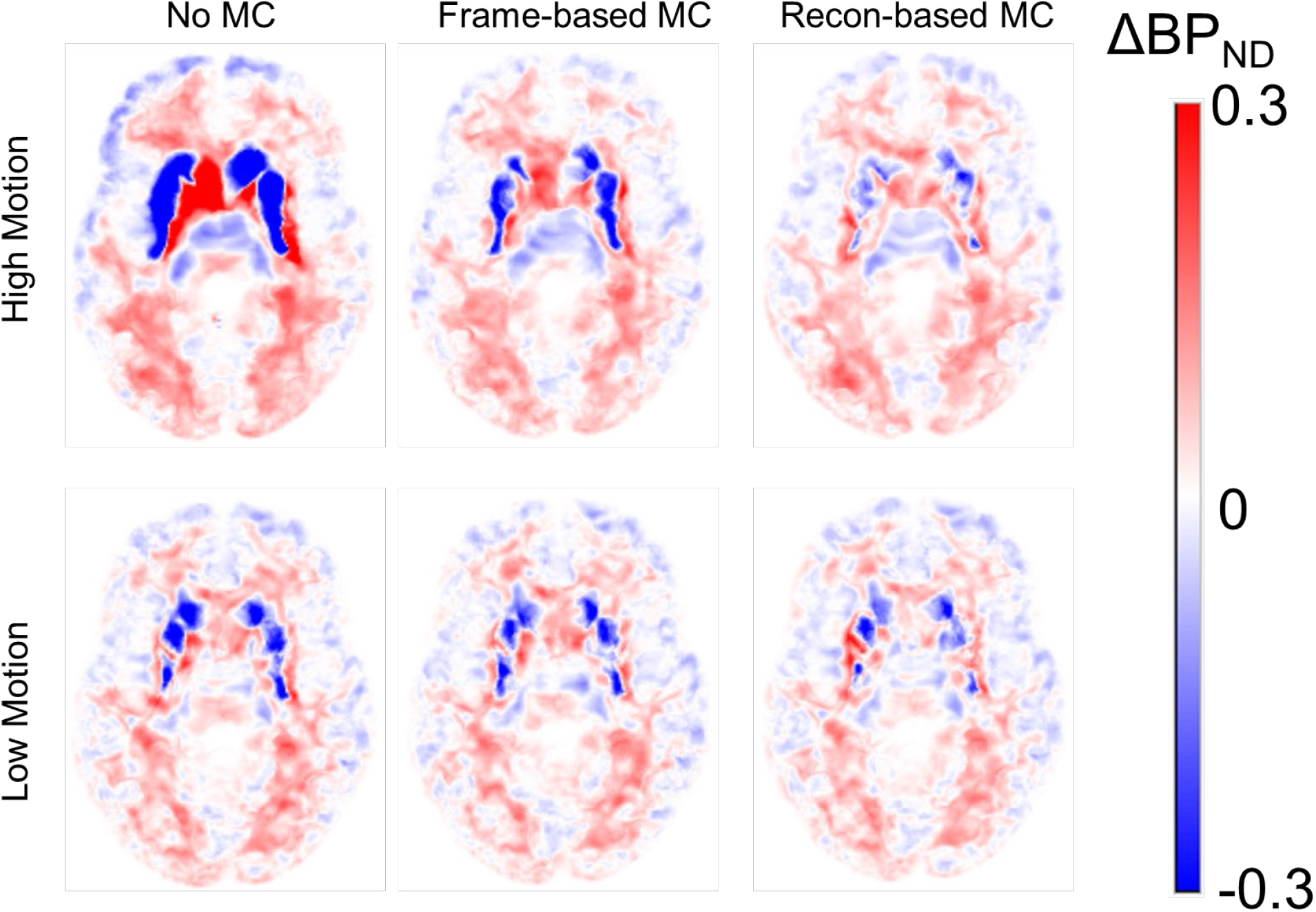
Motion Correction vs. Task Response. Parametric maps of task response (ΔBP_ND_) fit with an extension of MRTM2 using different approaches for motion correction (no motion correction, frame-based motion correction, reconstruction based motion correction) averaged across participants within the high (n = 10) and low (n = 10) motion groups.

**Table 1:** Regional averages of binding potential and task response. Average and SD values are averaged across participants within the high and low motion groups.

| High | Motion Correction | BP<sub>ND</sub> | BP<sub>ND</sub> SD | ΔBP<sub>ND</sub> | ΔBP<sub>ND</sub> SD | %ΔBP<sub>ND</sub> |
| --- | --- | --- | --- | --- | --- | --- |
| Putamen | None | 3.30 | 0.45 | -0.35 | 0.64 | -10.7% |
|  | Frame | 3.15 | 0.26 | -0.09 | 0.06 | -2.9% |
|  | Reconstruction | 3.16 | 0.22 | -0.02 | 0.06 | -0.5% |
| Caudate | None | 1.64 | 2.39 | 0.20 | 2.31 | 12.0% |
|  | Frame | 1.75 | 0.30 | -0.04 | 0.11 | -2.2% |
|  | Reconstruction | 2.07 | 0.17 | -0.03 | 0.05 | -1.5% |
| Nucleus Accumbens | None | 2.26 | 0.23 | -0.28 | 0.57 | -12.2% |
|  | Frame | 2.23 | 0.38 | -0.03 | 0.08 | -1.3% |
|  | Reconstruction | 2.05 | 0.27 | 0.04 | 0.06 | 1.9% |
| Low | Motion Correction | BP<sub>ND</sub> | BP<sub>ND</sub> SD | ΔBP<sub>ND</sub> | ΔBP<sub>ND</sub> SD | %ΔBP<sub>ND</sub> |
| Putamen | None | 3.61 | 0.38 | -0.10 | 0.06 | -2.7% |
|  | Frame | 3.50 | 0.40 | -0.07 | 0.05 | -2.0% |
|  | Reconstruction | 3.44 | 0.38 | -0.01 | 0.03 | -0.3% |
| Caudate | None | 2.17 | 0.24 | -0.14 | 0.12 | -6.2% |
|  | Frame | 1.85 | 0.28 | -0.08 | 0.06 | -4.6% |
|  | Reconstruction | 2.05 | 0.25 | -0.09 | 0.05 | -4.4% |
| Nucleus Accumbens | None | 2.41 | 0.36 | -0.06 | 0.23 | -2.4% |
|  | Frame | 2.48 | 0.37 | -0.06 | 0.11 | -2.6% |
|  | Reconstruction | 2.29 | 0.32 | 0.04 | 0.10 | 1.7% |

### Simulating Motion Bias

Figure 7. explores the impact of motion bias at different levels of task response. When there is no task response (0%), motion bias can make it appear as though there is a modest (ΔBP_ND_ = −3.1% ± 2.2%) response. However, at high magnitudes of task response (20%), the error in the overall observed effect (ΔBP_ND_ = −24.5% ± 7.3%) is attributable primarily to model bias with motion bias playing a much smaller role (Table 2).

**Figure 7:**
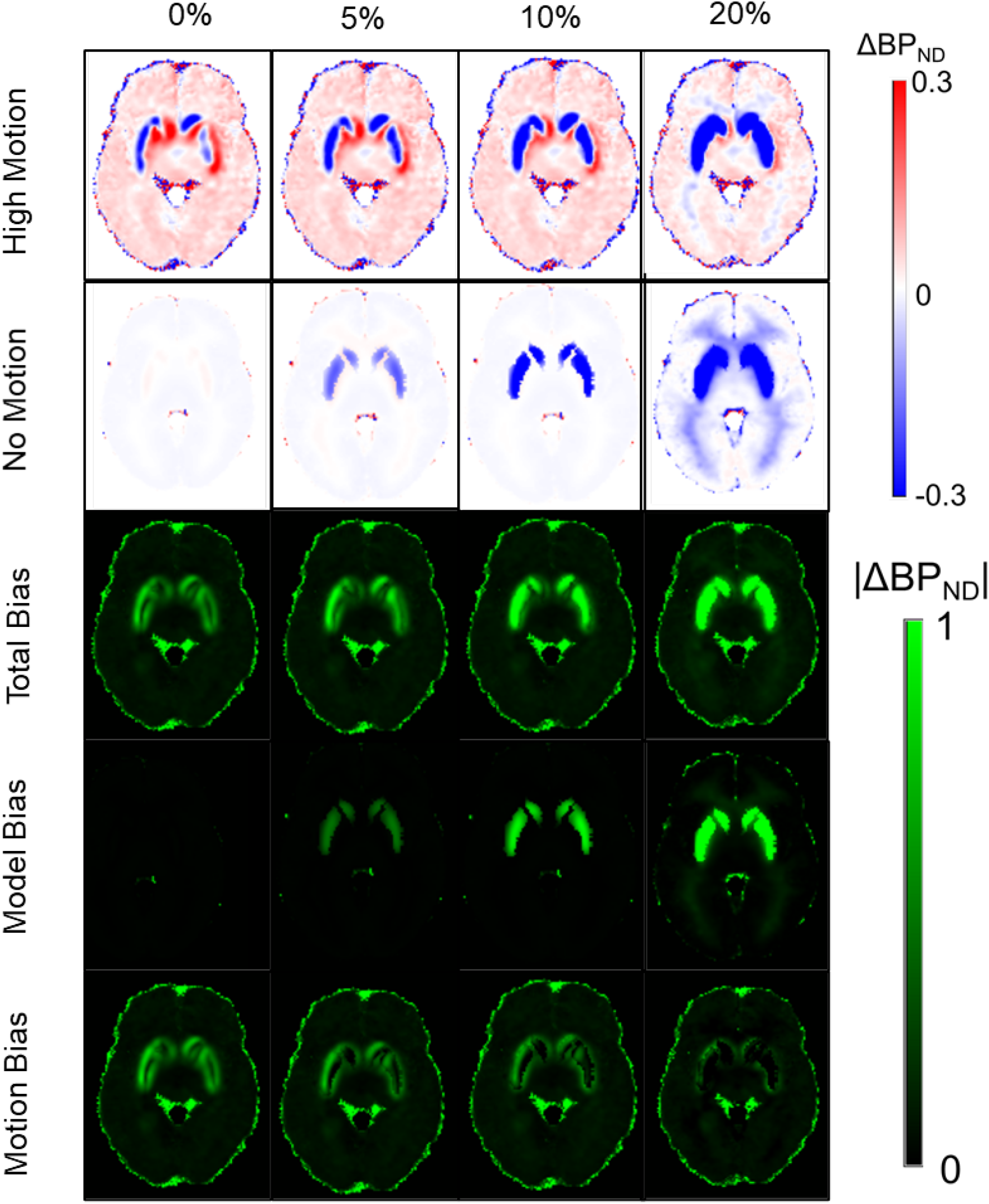
Motion bias simulation experiment. Each column depicts the simulation of a different level of challenge response in terms of change in BP_ND_. The top two rows depict the ΔBP_ND_ levels in high motion and motionless simulations respectively. The bottom three rows show the absolute value of the error in ΔBP_ND_ relative to the simulated response magnitude for each column. Total bias is derived from the high motion simulations, model bias from the motionless simulations, and motion bias from the difference between them.

**Table 2:** Simulated absolute value of parametric ΔBP_ND_ bias levels in putamen (**Figure 7**). Total bias was derived from the high motion simulations, model bias from the motionless simulations, and motion bias from the difference between them. Values are µ ± SEM. Noiseless estimates of model bias do not have associated error estimates.

| Challenge Response | BPnd | ΔBPnd | %ΔBPnd | \|%ΔBPnd Error\|<br>Total Bias | \|%ΔBPnd Error\|<br>Model Bias | \|%ΔBPnd Error\|<br>Motion Bias |
| --- | --- | --- | --- | --- | --- | --- |
| 0% | $3.6 \pm 0.07$ | $-0.11 \pm 0.08$ | $-3.1 \pm 2.2$ | $17.5\% \pm 3.7$ | 0.2% | $17.3\% \pm 3.7$ |
| 5% | $3.6 \pm 0.07$ | $-0.23 \pm 0.09$ | $-6.3 \pm 2.5$ | $20.6\% \pm 2.7$ | 8.0% | $12.3\% \pm 2.7$ |
| 10% | $3.6 \pm 0.07$ | $-0.39 \pm 0.10$ | $-11.0 \pm 2.8$ | $26.4\% \pm 2.5$ | 17.4% | $9.0\% \pm 2.5$ |
| 20% | $3.7 \pm 0.16$ | $-0.92 \pm 0.27$ | $-24.5 \pm 7.3$ | $46.4\% \pm 7.7$ | 48.6% | $0.0\% \pm 5.5$ |

### Simulating Intraframe Motion

Figure 8. depicts the differential impact on ΔBP_ND_ of simulated intraframe versus total motion averaged across realizations of estimates from the high motion group. With no simulated challenge, all of the measured change in BP_ND_ can be attributed to bias: the combined effect of inter- and intraframe motion. After frame-based motion correction, the remaining bias attributable to intraframe motion was greatly reduced.

**Figure 8:**
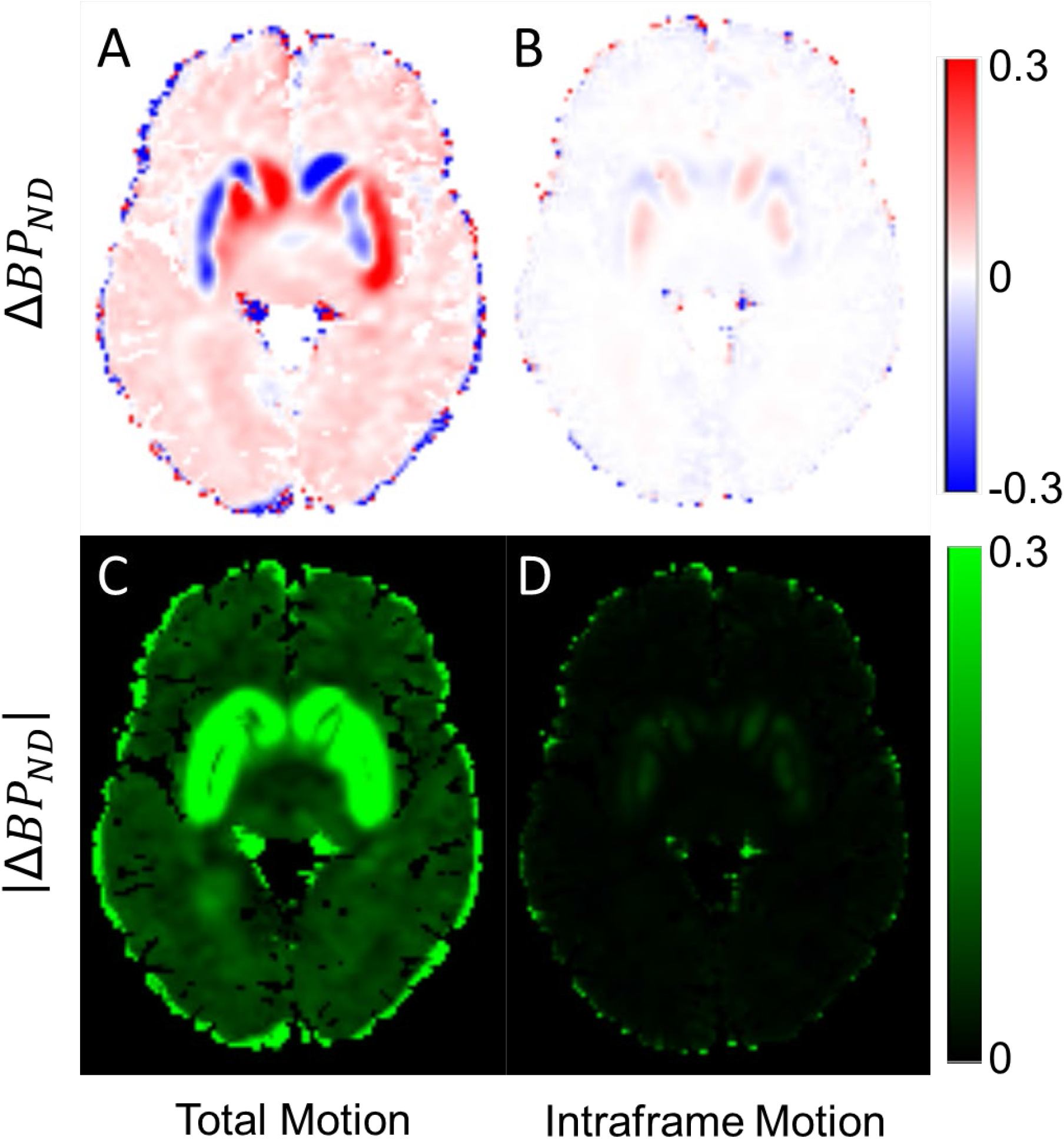
Simulated effect of total motion vs. intraframe motion averaged across realizations. (A) Total motion bias in ΔBP_ND_. (B) Motion bias in ΔBP_ND_ after frame-based motion correction. (C) Absolute value of total motion bias in ΔBP_ND_. (D) Absolute value of motion bias in ΔBP_ND_ after frame-based motion correction.

With both positive and negative motion bias apparent at the voxel level, absolute bias values were taken at the regional level to avoid underestimating the total effect magnitude. Without performing motion correction, the absolute value of the total motion bias in putamen ΔBP_ND_ was 0.63 ± 0.12 standard error (SE). After frame-based motion correction, the remaining absolute bias attributable to intraframe motion was 0.048 ± 0.003 SE (a drop in motion bias of 92.4% ± 5.6% SE). For comparison, putamen RAC BP_ND_ has been shown to have a standard deviation of 10% in test-retest evaluations^52^.

## DISCUSSION

An examination of motion bias has been presented in the context of an intrascan behavioral challenge response paradigm. While motion bias was observed to be of comparable magnitude to the task response, it was also mostly controllable using frame-based motion correction. Starting from this expected result, we were able to pose more detailed questions such as precisely how much finer-grained motion estimation and correction reduces bias, how responses of different magnitudes are affected by a high but plausible degree of motion bias, and how much intraframe motion contributes to bias relative to the total motion.

RAC studies are especially vulnerable to motion corruption because tracer uptake is highly localized to the striatum, with steep gradients in binding potential in the surrounding areas. Therefore, any unresolved motion will result in spillover of activity signal (and therefore binding potential) into the voxels immediately surrounding the striatum. This is especially problematic for studies with intra-scan challenges because such changes in binding potential are precisely what the additional challenge term in the kinetic model is intended to fit. Uncorrected motion between PET frames will therefore create artifacts showing decreases in binding potential on one side of the striatum and matching increases on the opposite side (**Figure 6**).

Simulating PET data with different levels of task response, we were able to separate the true task response effect from motion bias. In **Figure 7**, we can see how the relative contribution of motion bias to the total effect decreases for higher task responses. At 0% simulated response, large positive and negative biases in ΔBP_ND_ predominate, meeting in the center of the striatum. As the response level increases however, the measured ΔBP_ND_ in the striatum becomes more reliably negative, as the greater dopamine D_2_/D_3_ challenge response begins to outweigh the motion bias. Another effect worth noting is the increase in model bias associated with increasing task response. Though we prevented one form of model bias by both simulating and fitting a 1TCM topology, we introduced another by fixing k_2_’ and reducing the model from 4 parameters to 3. This is the primary tradeoff of the SRTM2 framework: decreasing noise at the expense of bias^46^. As MRTM tends to underestimate k_2_’^53^, fixing an underestimated value of k_2_’ for use with MRTM2 can also lead to biased estimates of ΔBP_ND_^42^, which is the source of bias that comes to predominate at higher task response magnitudes.

As the biggest difference between the standard frame-based motion correction and other more advanced forms of motion correction is their capacity to correct for intraframe motion, we used simulations to determine the relative contribution of intra- and interframe motion to bias in ΔBP_ND_.

Figure 8. shows that interframe motion contributes the majority of the bias effect. This is likely because of how each affects the TACs that make up the PET images. Fundamentally, interframe motion is a displacement, with the striatum moving from one position to another from one frame to the next. This creates TACs with discontinuous or stepped appearances, which are liable to be fit as changes in binding potential. Once motion between the frames has been accounted for, intraframe motion appears as a spatial blurring without displacement of the overall frame. Spatial blurring alone is unlikely to significantly bias voxelwise estimates of ΔBP_ND_. In fact, Gaussian smoothing is often applied to PET images prior to kinetic modeling to reduce noise. The residual motion bias in ΔBP_ND_ after frame-based motion correction can just as easily be explained by the limits of motion estimation/registration accuracy. These results indicate that even PET studies with modest response magnitudes can be effectively motion corrected using a standard frame-based approach.

Our study had several limitations. The observed bias pattern is specific to RAC and may not be as significant to other tracers which have neighboring regions more similar in their activity levels. Furthermore, the simulations did not consider the potential effects of attenuation-emission mismatches, though others have shown that such effects are not especially pronounced for RAC^33^. Another limitation of our study is that its high dose and bolus plus constant infusion paradigm may not be representative of most RAC studies. With its highly localized uptake making RAC more difficult to register than a radiotracer that is more generally distributed throughout the brain, such as ^18^F-fluorodeoxyglucose, similar studies with lower dose may not be able to rely on either frame-based correction or PET data driven motion estimation. Such cases would instead require sacrificing temporal resolution in PET-based estimates (i.e., by using longer frames when counts are lower) or else increasing complexity by estimating motion using another simultaneously acquired source (i.e., MR, optical). Finally, the regional results reported in Table 1 are limited by the spatial variation in BP_ND_ and ΔBP_ND_ within a given region. For example, in **Figure 6**, the greatest areas of negative ΔBP_ND_ clearly correspond to the subcortical regions of the striatum. However, the edges that divide the positive from the negative ΔBP_ND_ values lie within the area defined by the anatomically derived region of interest labels, creating average regional measurements that are more liable to miss localized task responses.

In conclusion, the effects of motion bias on behavioral task response have been investigated and characterized in the context of an intrascan RAC fMRI challenge study, demonstrating that the impact of uncorrected motion bias can be commensurate with that of the challenge response and can even account for the majority of the apparent effect at the regional level. This work highlights the importance of carefully accounting and correcting for motion, which is essential for fully realizing the multimodality benefits of simultaneous RAC PET and fMRI in behavioral challenge experiments.

## Data Availability

Please contact the corresponding author for any inquiries into data availability.

## ACKNOWLEDGMENTS

We would like to thank Rajesh Narendran for making available to us a set of previously published RAC data with arterial blood sampling^5^ for comparison.

This work was partly supported by National Institute of Biomedical Imaging and Bioengineering Grant 5R01EB014894-02, National Institute of Mental Health Grant Number R01MH080243, National Institute of General Medical Sciences Grant T32 GM008313, NIH Blueprint for Research Science Grant T90DA022759/R90DA023427, and NIH Shared Instrumentation Grant S10RR023043.

